# Brain age gap correlates with DTI-derived microstructural abnormalities in multiple sclerosis

**DOI:** 10.64898/2026.06.15.26355725

**Authors:** Stasson Lea, Oun Al-iedani, Caden Bardell, Sean Burnard, Vicki Maltby, Saad Ramadan, Rodney Lea, Jeannette Lechner-Scott

**Author notes:** Joint last authors (Rodney Lea, Jeannette Lechner-Scott). Corresponding Author: Rodney Lea.

## Abstract

**Background:** Brain age gap (BAG) is increased in multiple sclerosis (MS), but whether it reflects microstructural pathology beyond conventional atrophy remains unclear.

**Objective:** To test whether BAG is elevated in MS and correlates with conventional and diffusion tensor imaging (DTI) abnormalities relative to healthy controls.

**Methods:** A case-control study of 43 people with MS and 18 healthy controls was performed. BAG was estimated from T1-weighted MRI using brainageR. Controls were used as MRI reference distributions. MRI values were expressed as deviation z-scores and correlated with BAG within MS. Conventional MRI and DTI domains were analysed using age/sex-adjusted partial correlations with domain-wise Benjamini-Hochberg FDR correction, where appropriate.

**Results:** BAG was higher in MS than controls (4.79 vs −2.58 years; p<0.001; Cohen’s d=0.84). Within MS, BAG correlated with EDSS (partial r=0.38, p=0.014), disease duration (r=0.39, p=0.011), and lesion volume (r=0.67, p<0.001). Control-referenced conventional MRI abnormalities correlated strongly with BAG, including lower peripheral grey matter volume (r=−0.71, q<0.001), higher CSF volume (r=0.69, q<0.001), and lower grey matter volume (r=−0.67, q<0.001). DTI associations were robust, including higher NAWM mean diffusivity (r=0.66, q<0.001), higher radial diffusivity (r=0.65, q<0.001), and lower fractional anisotropy (r=−0.52, q<0.001).

**Conclusions:** BAG was elevated in MS and correlated with clinical severity, conventional MRI abnormality, and DTI-derived microstructural injury. These findings support BAG as a biologically relevant MS phenotype extending beyond volumetric atrophy.

## INTRODUCTION

Multiple sclerosis (MS) is characterised by cerebral demyelination, axonal injury and neurodegeneration. Conventional MRI remains central to MS diagnosis and monitoring, but standard clinical metrics incompletely capture disease burden. Lesion measures index focal inflammatory injury, while volumetric measures reflect accumulated tissue loss. However, these markers do not fully represent diffuse normal-appearing white matter injury or network-level vulnerability relevant to disability and cognitive decline [14–18].

Brain age modelling provides an imaging-based estimate of deviation from expected healthy ageing. Brain age gap (BAG), calculated as MRI-estimated brain age minus chronological age, is consistently increased in MS and has been associated with disability, lesion burden, brain atrophy, cognitive performance, and future disability accumulation [1–11]. These findings support the clinical relevance of BAG, but its biological interpretation in MS remains incompletely resolved.

Most MS brain-age estimates are derived from T1-weighted structural MRI or volumetric and morphological features. The brainageR package, for example, estimates brain age from raw T1-weighted MRI using SPM12 segmentation and normalisation followed by Gaussian process regression [13]. Consequently, associations between BAG and conventional T1-derived volumetric measures may partly reflect shared imaging information rather than independent biological validation. This limitation is increasingly recognised, with recent work emphasising disease-specific modelling, lesion handling, preprocessing harmonisation, and interpretability [5,6,10,11].

A key next step is to test whether BAG is associated with MRI measures that are biologically relevant to MS but less directly embedded in the T1-derived brain-age signal. Diffusion tensor imaging (DTI) quantifies white matter microstructure through fractional anisotropy, mean diffusivity, axial diffusivity, and radial diffusivity, and is sensitive to pathology in lesions and normal-appearing white matter [17,18].

Here, we examined BAG in an MRI dataset comprising conventional volumetrics, lesion burden, and DTI. Previous analyses showed that multimodal MRI signatures predicted five-year cognitive decline more strongly than volumetric measures alone, with informative features spanning DTI and volumetric domains [12]. In the present cross-sectional case-control analysis, we first tested whether BAG was elevated in MS relative to healthy controls. We then tested, within MS, whether BAG correlated with conventional and DTI-derived MRI abnormalities expressed relative to healthy-control reference values. We hypothesised that higher BAG would be associated not only with conventional volumetric and lesion measures, but also with DTI measures of white matter microstructure, supporting BAG as a biologically anchored MRI phenotype rather than a surrogate for brain atrophy alone.

## RESULTS

### Participant characteristics

The baseline analytic study included 61 participants, comprising 43 people with MS and 18 healthy controls. Sex distribution was similar between groups, with 31 of 43 MS participants (72.1%) and 13 of 18 controls (72.2%) being female. Mean age was also similar between MS participants and controls (43.5 vs 43.0 years). In the MS group, median Expanded Disability Status Scale (EDSS) score was 2.0, mean disease duration was 7.47 years, mean age at onset was 36.0 years, mean Symbol Digit Modalities Test (SDMT) score was 51.2, mean total Audio Recorded Cognitive Screen (tARCS) score was 86.6 and mean Modified Fatigue Impact Scale (MFIS) score was 31.5. Compared with healthy controls, MS participants had lower SDMT scores and higher fatigue and mood symptom scores; tARCS was numerically lower but did not meet conventional statistical significance.

**Table 1.** Baseline demographic and clinical characteristics of study participants.

| Characteristic | MS (n=43) | Healthy controls (n=18) | p value |
| --- | --- | --- | --- |
| Participants, n | 43 | 18 | — |
| Female sex, n (%) | 31 (72.1) | 13 (72.2) | 1.000 |
| Age, years | 43.5 ± 10.7 | 43.0 ± 10.2 | 0.868 |
| Brain age gap, years | 4.8 ± 9.8 | -2.6 ± 5.4 | 0.00045 |
| EDSS, median (IQR) | 2.0 (1.5–2.3) | — | — |
| Disease duration, years | 7.5 ± 6.0 | — | — |
| Age at onset, years | 36.0 ± 9.5 | — | — |
| Total lesion volume, median (IQR) | 3.2 (1.6–6.4) | — | — |
| Dimethyl fumarate, n (%) | 9 (20.9) | — | — |
| Fingolimod, n (%) | 21 (48.8) | — | — |
| Injectable therapy, n (%) | 13 (30.2) | — | — |
| SDMT | 51.2 ± 9.6 | 59.6 ± 16.1 | 0.026 |
| tARCS | 86.6 ± 16.1 | 94.9 ± 15.7 | 0.070 |
| MFIS | 31.5 ± 18.6 | 15.6 ± 11.3 | 0.00015 |
| DASS-21 total | 25.3 ± 25.4 | 8.9 ± 8.8 | 0.00041 |
| DASS-21 stress | 11.4 ± 10.8 | 6.1 ± 6.7 | 0.024 |
| DASS-21 anxiety | 6.7 ± 7.2 | 1.7 ± 2.5 | 0.00014 |
| DASS-21 depression | 7.5 ± 9.4 | 1.1 ± 1.6 | 0.000086 |

Values are mean ± SD unless otherwise specified. EDSS and total lesion volume are reported as median (IQR). p values compare MS and healthy controls where variables were available in both groups; Welch's t-test was used for continuous variables and Fisher's exact test for sex. Treatment categories are reported for MS participants only. BAG, brain age gap; DASS-21, Depression Anxiety Stress Scales-21; EDSS, Expanded Disability Status Scale; IQR, interquartile range; MFIS, Modified Fatigue Impact Scale; SDMT, Symbol Digit Modalities Test; tARCS, total Audio Recorded Cognitive Screen.

### BAG differences in MS versus controls

BAG differed between groups, with higher BAG in MS than controls. Mean BAG, defined as predicted brain age minus chronological age, was 4.79 years in the MS cohort and −2.58 years in healthy controls, corresponding to a mean difference of 7.37 years (Welch p<0.001; Cohen’s d=0.84). Thus, MS participants had higher BAG on average, while the local healthy-control group had a mean BAG below zero. In the age/sex-adjusted model, MS status was associated with a 7.45-year higher BAG (p=0.004). These results confirmed that the study reproduced the expected elevation of BAG in MS. Mean BAG was numerically similar across treatment subgroups, and there was no statistical evidence of a treatment-subgroup difference in BAG at baseline.

### Routine clinical correlations with BAG in MS

Within MS, observed BAG showed expected correlations with routine clinical indices. Higher BAG was associated with higher EDSS (partial r=0.38, p=0.014), longer disease duration (partial r=0.39, p=0.011), lower derived age at onset (partial r=−0.39, p=0.011), and greater log-transformed lesion volume (partial r=0.67, p<0.001). These associations provide internal consistency with prior MS brain-age studies reporting higher BAG in relation to disability, disease burden, lesion load and progression-related outcomes [1–4,11]. The association with derived age at onset should be interpreted cautiously because age at onset was calculated from chronological age and disease duration.

### Conventional MRI correlations with BAG in MS

Conventional MRI measures were first normalised to healthy controls and age/sex-adjusted before correlation with BAG within MS. For each MRI metric, healthy controls were used as an internal reference group by fitting an age- and sex-adjusted model in controls and expressing MS values as residual z-scores relative to the control residual distribution. Because the healthy-control group was modest, these values should be interpreted as internal deviation scores rather than definitive normative centiles. As shown in Figure 3, the direction of association was consistent with greater tissue loss in participants with higher BAG: lower peripheral grey matter volume (partial r=−0.71, p<0.001, domain-wise FDR q<0.001), lower grey matter volume (partial r=−0.67, p<0.001, q<0.001), lower total brain volume (partial r=−0.58, p<0.001, q<0.001), lower white matter volume (partial r=−0.33, p=0.032, q=0.032), and higher CSF volume (partial r=0.69, p<0.001, q<0.001). These associations survived domain-wise FDR correction.

**Figure 1.**
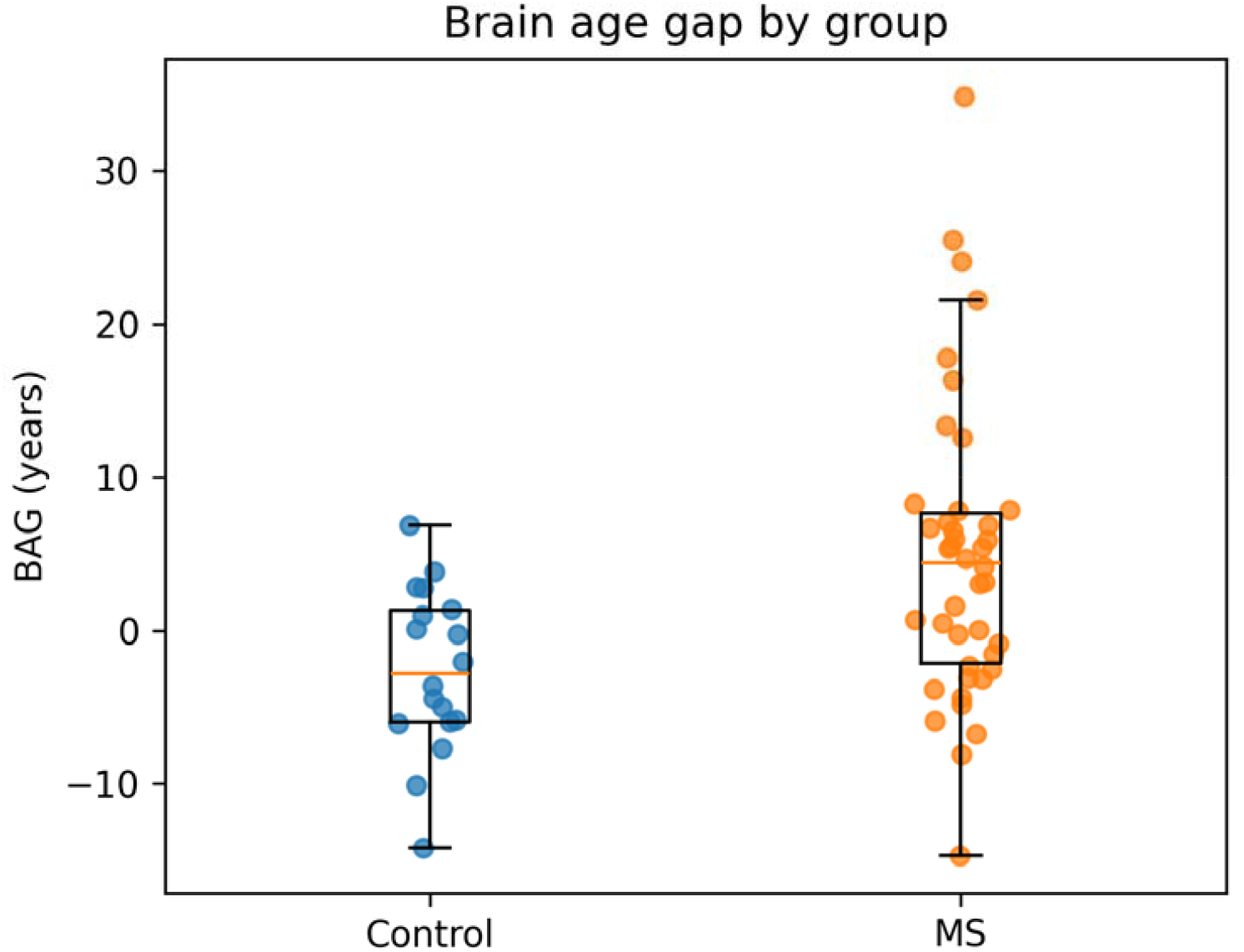
BAG distribution in MS and healthy controls. Boxplots with participant-level points show observed BAG in years by group

**Figure 2.**
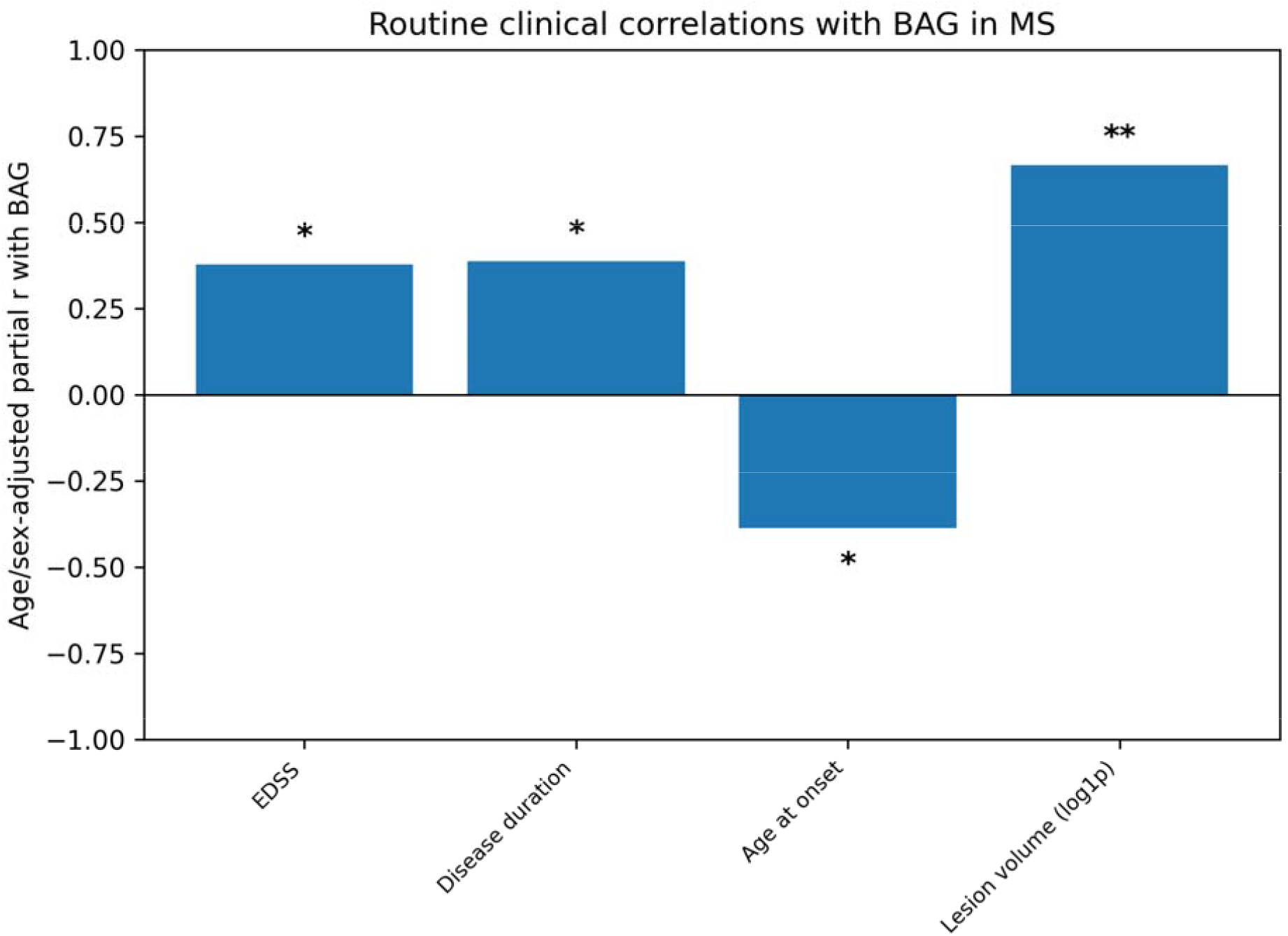
Routine clinical correlations with BAG in MS. Bars show age/sex-adjusted partial r. Asterisks indicate raw p-value thresholds: *p<0.05 and **p<0.005.

**Figure 3.**
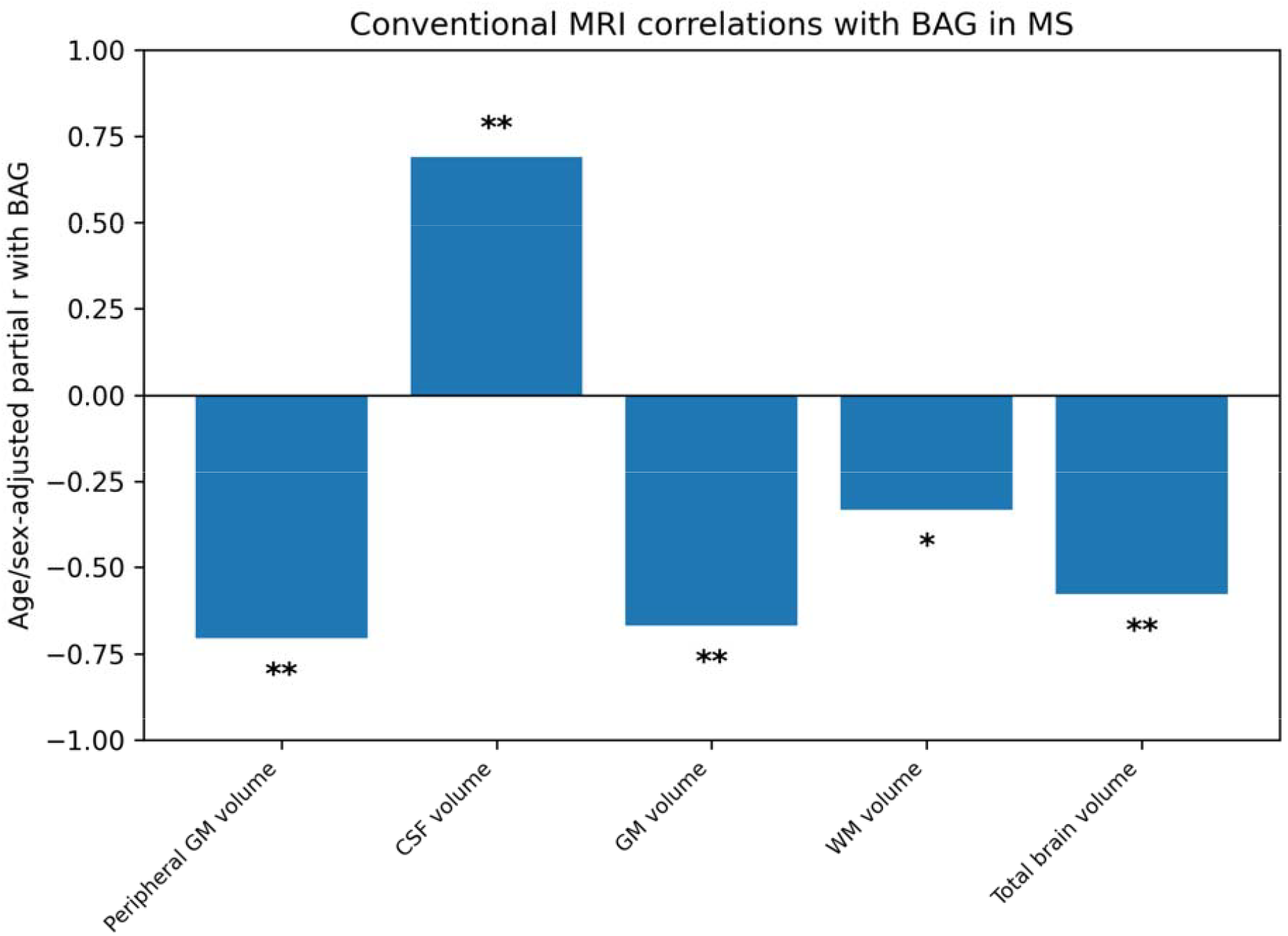
Conventional MRI correlations with BAG in MS. MRI values were expressed as healthy-control-referenced age/sex-adjusted deviation z-scores before correlation within MS. Bars show age/sex-adjusted partial r. Asterisks indicate domain-wise BH-FDR thresholds: *q<0.05 and **q<0.005.

**Figure 4.**
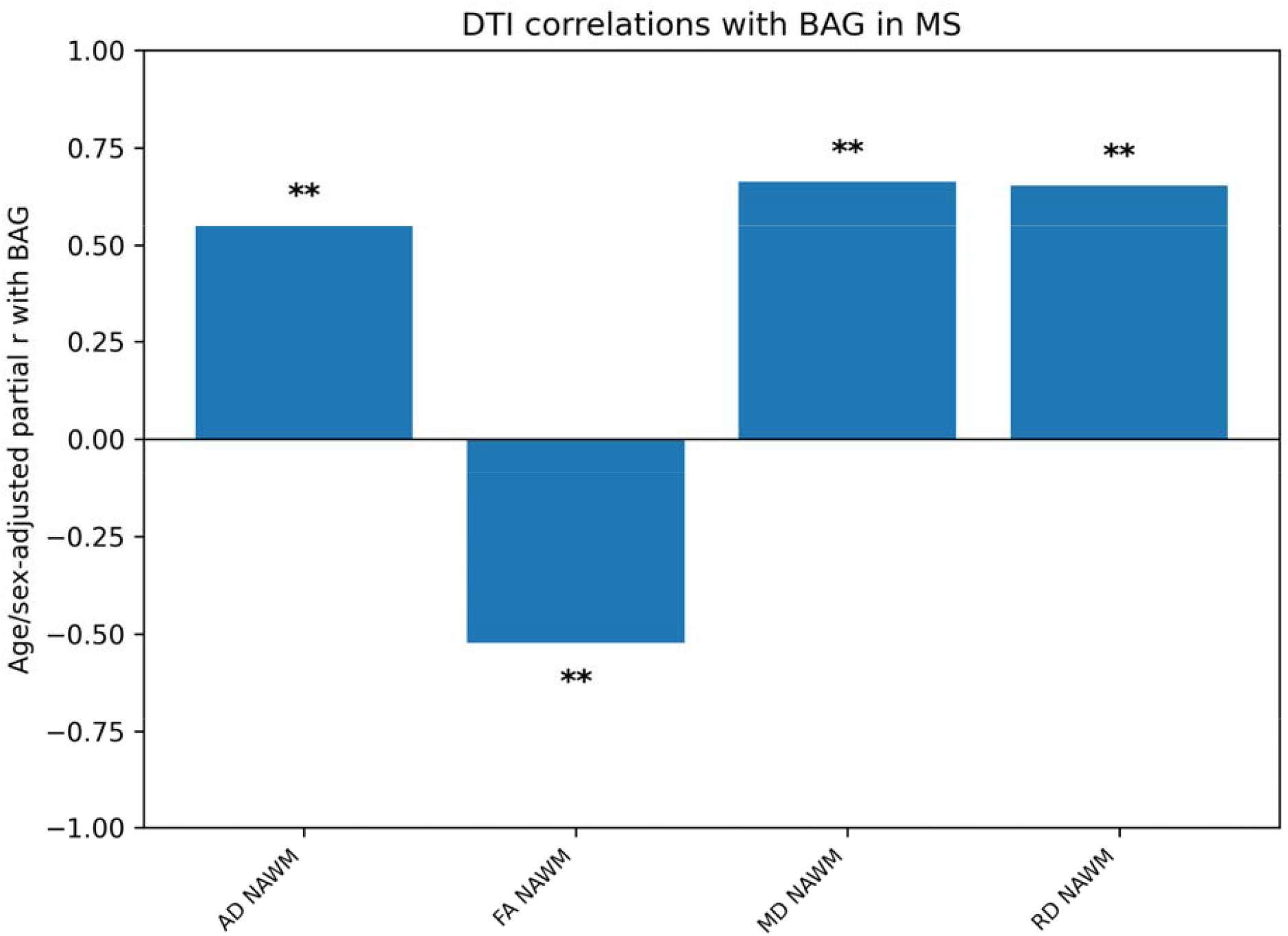
DTI correlations with BAG in MS. DTI values were expressed as healthy-control-referenced age/sex-adjusted deviation z-scores before correlation within MS. Bars show age/sex-adjusted partial r. Asterisks indicate domain-wise BH-FDR thresholds: *q<0.05 and **q<0.005.

**Figure 5.**
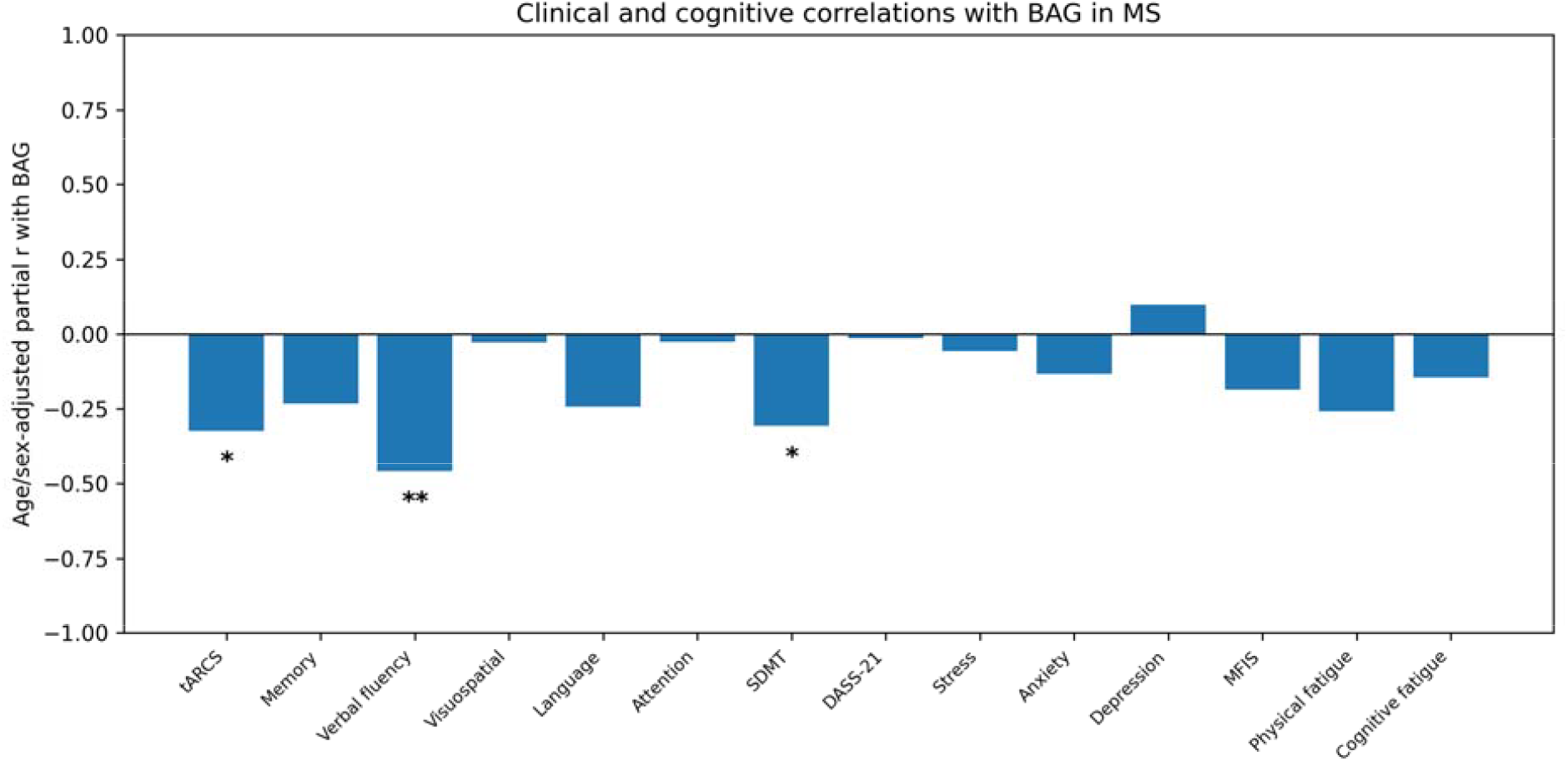
Clinical and cognitive correlations with BAG in MS. Bars show age/sex-adjusted partial r. Asterisks indicate raw p-value thresholds: *p<0.05 and **p<0.005.

### DTI correlations with BAG in MS

DTI analyses included 40 MS participants with referenceable DTI data. Three MS participants were not included in DTI analyses because DTI data were unavailable or unsuitable for the reference-based DTI workflow. Higher BAG was associated with higher normal-appearing white matter mean diffusivity (partial r=0.66, p<0.001, domain-wise FDR q<0.001), higher radial diffusivity (partial r=0.65, p<0.001, FDR q<0.001), higher axial diffusivity (partial r=0.55, p<0.001, FDR q<0.001) and lower fractional anisotropy (partial r=−0.52, p<0.001, FDR q<0.001). These directions were consistent with reduced microstructural white-matter integrity in participants with older appearing brains. The figure provides the domain-level pattern.

### Other clinical correlations in MS

Exploratory clinical and cognitive analyses were performed in MS cases only (Figure 5). These analyses included SDMT, tARCS and ARCS subdomains, fatigue and mood measures, excluding the routine clinical variables already reported above and shown in Figure 2. Results were interpreted as exploratory using raw p-values and were not used to define the primary imaging conclusions.

A full table of correlation statistics with BAG in MS is shown in Supplementary File 1.

## DISCUSSION

In this cross-sectional case-control study, BAG was elevated in MS and correlated with routine clinical indices and MRI abnormalities within MS. Mean BAG, defined as predicted brain age minus chronological age, was 4.79 years in MS compared with −2.58 years in controls, indicating that MS participants had higher BAG than local healthy controls. Within MS, higher BAG was associated with greater disability, longer disease duration, lower age at onset and higher lesion burden. The main imaging analyses showed that BAG correlated with conventional MRI abnormalities. The novel finding is that DTI measures of normal-appearing white matter structure also correlated with BAG after accounting for healthy-control reference values.

These findings extend prior MS brain-age literature in a biologically important way. Previous studies have established that people with MS show elevated BAG and that BAG relates to disability, lesion burden, brain parenchymal fraction, and clinical progression. However, much of this evidence has relied on structural or volumetric MRI, which is close to the imaging information used to derive many brain-age estimates. The present results support the interpretation that BAG is not only a T1-derived volumetric ageing index but is also linked to broader MS-relevant microstructural abnormalities.

### Relationship to existing MS brain-age literature

The observation of elevated BAG in MS was directionally consistent with previous reports showing that people with MS have older-appearing brains than age-matched controls. Recent reviews have summarised MS brain-age studies as showing average BAG increases of approximately 5-15 years, with older-appearing brains linked to disability, lesion burden, reduced total brain volume, cognitive outcomes, and future progression. Longitudinal and real-world studies have supported the prognostic relevance of brain age for disability accumulation, while cross-sectional cognitive studies have linked higher BAG with lower cognitive performance [11].

The present study did not simply replicate these findings. Its main contribution was to test whether BAG was coupled to MRI abnormalities that were not conventional brain-age validation metrics. This distinction matters because brain-age estimates derived from T1-weighted MRI are expected to correlate with atrophy-related measures. By showing that control-referenced DTI abnormalities correlated with BAG within MS, the present analysis provided biological support for BAG as an MRI phenotype extending beyond conventional volumetric abnormality.

### DTI and microstructural anchoring of BAG

The DTI findings are central to the interpretation of BAG as more than a macrostructural atrophy marker. BAG correlated strongly with normal-appearing white matter diffusion metrics, including higher mean, radial and axial diffusivity and lower fractional anisotropy. This result is biologically plausible because diffusion MRI detects abnormalities in tissue microstructure, including normal-appearing white matter injury that is not fully visible on conventional MRI [18]. Diffusion abnormalities in MS may reflect demyelination, axonal damage, oedema, inflammation, gliosis or tissue disorganisation.

The direction and magnitude of these associations support the view that higher BAG in MS may reflect cumulative microstructural tissue injury, including axonal and myelin-related disruption, rather than only visible lesion burden or global atrophy. The finding also aligns with broader neuroimaging work suggesting that diffusion MRI captures ageing-relevant microstructural changes that may precede or complement macrostructural change.

### Directionality and pathological coherence

The direction of significant associations was consistent with expected MS pathology. Higher BAG was associated with lower tissue volumes, higher CSF volume and higher lesion volume, consistent with atrophy and lesion burden. Higher BAG was also associated with higher normal-appearing white matter diffusivity and lower fractional anisotropy, consistent with reduced microstructural integrity. This directional coherence strengthens the interpretation that BAG reflects biologically meaningful MS-related tissue injury rather than nonspecific statistical association.

### Clinical and translational implications

These findings support BAG as a biologically important MRI phenotype in MS, but not yet as a clinically deployable biomarker. The appeal of BAG is that it provides a single intuitive summary of diffuse brain injury, potentially useful for communicating cumulative disease burden and stratifying risk in future studies. If BAG were associated only with volumetric measures, its incremental biological value over established atrophy markers would be limited. By linking BAG to control-referenced DTI abnormalities, this study suggests that BAG may index a broader injury state involving microstructural disruption. Clinical utility will require prospective validation against longitudinal outcomes, treatment response, and independent cohorts.

### Limitations

Several limitations require consideration. First, the analysis was cross-sectional and therefore could not establish whether BAG predicted subsequent disability, cognitive decline, or future MRI change. Second, the study size was modest, particularly for healthy controls, limiting precision for control-referenced estimates; these values should therefore be interpreted as internal deviation scores rather than definitive normative centiles. Third, BAG was derived from T1-weighted MRI; therefore, associations with macrostructural measures may partly reflect shared imaging information. The DTI analyses were included specifically to address this issue, but external replication is required. Fourth, the findings were generated from a single cohort with a specific MRI acquisition protocol and require validation across scanners, processing pipelines, and independent MS populations.

## CONCLUSIONS

BAG was elevated in MS and correlated with routine clinical indices, conventional macrostructural MRI abnormalities, and DTI-derived microstructural injury metrics within MS. BAG was derived from the pretrained brainageR model as predicted brain age minus chronological age and was not re-centred to the local controls. In contrast, the MRI predictor variables were expressed as age/sex-adjusted deviation scores relative to the local healthy-control group. These findings support BAG as a biologically anchored MRI phenotype in MS and provide a rationale for longitudinal validation in larger independent cohorts.

## METHODS

### Study design and participants

We performed a cross-sectional case-control analysis of baseline data from the previously described sample. The original study was a prospective multimodal neuroimaging study designed to identify MRI signatures associated with cognitive decline in MS. The present analysis used baseline data only and included people with clinically definite MS and healthy controls with available T1-weighted brain MRI and multimodal MRI measures. The MS sample was drawn from the previously reported 5-year cohort in which participants remained on stable disease-modifying therapy during follow-up. The analytic study included 61 participants: 43 people with MS and 18 healthy controls.

The original cohort design, recruitment, eligibility criteria and imaging protocol have been reported previously [12]. Participants with MS had clinically definite disease according to the McDonald diagnostic criteria [19,20], underwent neurological and cognitive assessment, and completed multimodal MRI on the same day as clinical assessment. The study was approved by the Hunter New England Local Health District Human Research Ethics Committee (approval number 2024/ETH02685) and all participants provided written informed consent.

### Clinical and demographic variables

The primary demographic covariates were chronological age at MRI and sex. Disease group was defined as MS case or healthy control. Routine MS clinical indices included EDSS, disease duration, age at onset derived as chronological age at MRI minus disease duration, and log-transformed total lesion volume. Additional clinical and cognitive variables, including Symbol Digit Modalities Test (SDMT), total Audio Recorded Cognitive Screen (tARCS) and ARCS subdomain scores, fatigue (Modified Fatigue Impact Scale; MFIS) and mood measures (Depression Anxiety Stress Scales; DASS), were analysed as exploratory correlates within MS cases.

### MRI acquisition and preprocessing

MRI data were acquired on a 3T Siemens Magnetom Prisma system with a 64-channel receiver head coil at the Hunter Medical Research Institute Imaging Centre. The imaging protocol included three-dimensional T1-weighted magnetisation-prepared rapid acquisition gradient-echo imaging, T2-weighted fluid-attenuated inversion recovery imaging, and diffusion-weighted MRI [12].

Structural MRI processing in the original multimodal MRI processing pipeline included brain extraction, segmentation, lesion quantification, and volumetric estimation using established tools including FSL and SPM-based methods. White matter lesion quantification used T2-FLAIR hyperintensities, lesion probability mapping, and lesion filling on T1-MPRAGE images. Diffusion MRI preprocessing included denoising, Gibbs ringing removal, susceptibility-induced and eddy-current correction, and bias-field correction, followed by estimation of fractional anisotropy, mean diffusivity, radial diffusivity, and axial diffusivity in normal-appearing white matter and white matter lesions [12].

### Brain age estimation

Brain-predicted age was estimated from T1-weighted MRI using the brainageR framework, which applies established brain-age prediction methods to raw T1-weighted MRI data [13]. BAG was defined as predicted brain age minus chronological age. The variable brain_PAD was used as the BAG outcome throughout the analysis. BAG remained in years and was not control-adjusted or standardised. BAG was retained in its original units as predicted brain age minus chronological age. The local healthy-control group had a mean BAG below zero, indicating that brainageR predicted their brain age to be younger than chronological age on average. Therefore, the primary case-control result should be interpreted as a relative elevation of BAG in MS compared with local controls, rather than as absolute calibration of brainageR to this sample. MRI metrics were converted to healthy-control-referenced age/sex-adjusted z-scores to quantify MRI abnormality relative to healthy reference values. These MRI deviation scores were then correlated with BAG within MS participants.

### MRI variable classes

MRI variables were organised into biologically interpretable MRI classes. The conventional MRI class included total peripheral grey matter, total CSF volume, total grey matter volume, total white matter volume and total brain volume. Total lesion volume was analysed with routine clinical indices and was not included in the conventional MRI figure. The DTI class included axial diffusivity, fractional anisotropy, mean diffusivity and radial diffusivity measured in normal-appearing white matter and white matter lesions; only referenceable DTI measures were included in the control-referenced MRI analyses. DTI analyses included 40 MS participants; three MS participants were excluded from DTI analyses because DTI data were unavailable or unsuitable for the reference-based DTI workflow. Constant or redundant variables were excluded before analysis.

### Statistical analysis

The analysis followed a clinically focused workflow. First, BAG was compared between MS cases and healthy controls using Welch’s t-test, supported by an age/sex-adjusted linear model. Cohen’s d was reported as a standardised case-control effect size.

Second, routine clinical correlations were calculated within MS cases between observed BAG and EDSS, disease duration, derived age at onset and log-transformed total lesion volume. These analyses were included as contextual, prior-supported clinical associations and used age/sex-adjusted partial correlations with raw p-values.

For MRI analyses, multiple-testing correction was performed within prespecified MRI measurement domains using Benjamini-Hochberg false discovery rate correction. Conventional MRI and DTI were treated as separate hypothesis families. Domain-wise FDR q<0.05 was considered statistically significant after correction within each MRI domain. For clinical and cognitive correlations, raw p-values were reported because routine clinical correlations were contextual and additional clinical/cognitive correlations were exploratory. Exact p-values were reported to three decimal places unless p<0.001.

## Author Contributions

Conceptualization: Stasson Lea, Rodney Lea, Jeannette Lechner-Scott. Methodology: Stasson Lea, Oun Al-iedani, Sean Burnard, Saad Ramadan, Rodney Lea, Jeannette Lechner-Scott. Formal analysis: Stasson Lea, Rodney Lea. Investigation and data curation: Stasson Lea, Oun Al-iedani, Caden Bardell, Sean Burnard, Saad Ramadan, Rodney Lea, Jeannette Lechner-Scott. Writing - original draft: Stasson Lea, Rodney Lea. Writing - review and editing: all authors. Supervision: Rodney Lea, Jeannette Lechner-Scott.

## Acknowledgements

The authors acknowledge the participants who contributed to this study and the clinical and imaging staff involved in data acquisition and study coordination. OA was supported by a Post Doctoral Fellowship from Multiple Sclerosis Australia.

## Declaration of Generative AI in the Writing Process

During preparation of this manuscript, the authors used ChatGPT to assist with language editing, formatting, and manuscript organisation. The authors reviewed and edited all AI-assisted outputs and take full responsibility for the content of the manuscript.

## Data Availability Statement

De-identified participant-level data are not publicly available because of participant consent and ethics restrictions. Aggregated analysis outputs supporting the reported results are provided in Supplementary File 1. Additional reasonable requests for analytic details may be directed to the corresponding author, subject to institutional governance and ethics approvals.

## Supplementary Methods

See Supplementary File 1. Full correlation statistical results

## REFERENCES

1. Cole JH, Raffel J, Friede T, Eshaghi A, Brownlee WJ, Chard D, et al. Longitudinal assessment of multiple sclerosis with the brain-age paradigm. Ann Neurol. 2020;88(1):93–105.

2. Hogestol EA, Kaufmann T, Nygaard GO, Beyer MK, Sowa P, Nordvik JE, et al. Cross-sectional and longitudinal MRI brain scans reveal accelerated brain aging in multiple sclerosis. Front Neurol. 2019;10:450.

3. Brier MR, Li Z, Ly M, Karim HT, Liang L, Du W, et al. Brain age predicts disability accumulation in multiple sclerosis. Ann Clin Transl Neurol. 2023;10(6):990–1001.

4. Denissen S, Engemann DA, De Cock A, Costers L, Baijot J, Laton J, et al. Brain age as a surrogate marker for cognitive performance in multiple sclerosis. Eur J Neurol. 2022;29(10):3039–3049.

5. Pontillo G, Prados F, Colman J, Kanber B, Abdel-Mannan O, Al-Araji S, et al. Disentangling neurodegeneration from aging in multiple sclerosis using deep learning: the brain-predicted disease duration gap. Neurology. 2024;103(10):e209976.

6. Skattebol L, Nygaard GO, Leonardsen EH, Kaufmann T, Moridi T, Stawiarz L, et al. Brain age in multiple sclerosis: a study with deep learning and traditional machine learning. Brain Commun. 2025;7(3):fcaf152.

7. Skattebol L, Lundby R, Overas MH, Sowa P, Celius EG, Harbo HF, et al. Quantitative susceptibility mapping of paramagnetic rim lesions in early multiple sclerosis: a cross-sectional study of brain age and disability. Neuroimage Rep. 2025;5(3):100277.

8. Romme CJA, Stanley EAM, Mouches P, Wilms M, Pike GB, Metz LM, et al. Analysis and visualization of the effect of multiple sclerosis on biological brain age. Front Neurol. 2024;15:1423485.

9. Chai L, Sun J, Zhuo Z, Wei R, Xu X, Duan Y, et al. Estimated brain age in healthy aging and across multiple neurological disorders. J Magn Reson Imaging. 2025;62(3):869–879.

10. Hannoun S, Fayad G, El Ayoubi NK, Khoury SJ. The effect of lesion filling on brain age estimation in multiple sclerosis. BMC Med Imaging. 2025;25(1):356.

11. Lea R, Lea S, Giovannoni G, Hawkes C, Levy M, Yeh EA, et al. Brain Age. Mult Scler Relat Disord. 2025;104:106891.

12. Al-iedani O, Lea S, Alshehri A, Maltby VE, Saugbjerg B, Ramadan S, et al. Multi-modal neuroimaging signatures predict cognitive decline in multiple sclerosis: a 5-year longitudinal study. Mult Scler Relat Disord. 2024;81:105379.

13. Cole JH, Poudel RPK, Tsagkrasoulis D, Caan MWA, Steves C, Spector TD, et al. Predicting brain age with deep learning from raw imaging data results in a reliable and heritable biomarker. NeuroImage. 2017;163:115–124. doi:10.1016/j.neuroimage.2017.07.059.

14. DeLuca GC, Yates RL, Beale H, Morrow SA. Cognitive impairment in multiple sclerosis: clinical, radiologic and pathologic insights. Brain Pathol. 2015;25(1):79–98.

15. Rocca MA, Amato MP, De Stefano N, Enzinger C, Geurts JJG, Penner IK, et al. Clinical and imaging assessment of cognitive dysfunction in multiple sclerosis. Lancet Neurol. 2015;14(3):302–317.

16. Pike AR, Branger P, De Stefano N, Stromillo ML, Giorgio A, et al. Neuroimaging predictors of longitudinal disability and cognition outcomes in multiple sclerosis patients: a systematic review and meta-analysis. Mult Scler Relat Disord. 2022;57:103452.

17. Andersen O, et al. Diffusion tensor imaging in multiple sclerosis at different final outcomes. Acta Neurol Scand. 2018;137(2):165–173.

18. Enzinger C, Barkhof F, Ciccarelli O, Filippi M, Kappos L, Rocca MA, et al. Nonconventional MRI and microstructural cerebral changes in multiple sclerosis. Nat Rev Neurol. 2015;11(12):676–686.

19. Polman CH, Reingold SC, Banwell B, Clanet M, Cohen JA, Filippi M, et al. Diagnostic criteria for multiple sclerosis: 2010 revisions to the McDonald criteria. Ann Neurol. 2011;69(2):292–302.

20. Thompson AJ, Banwell BL, Barkhof F, Carroll WM, Coetzee T, Comi G, et al. Diagnosis of multiple sclerosis: 2017 revisions of the McDonald criteria. Lancet Neurol. 2018;17(2):162–173.

